# Baseline characteristics of the population-based cohort in Iga City – the Iga City Cohort Study

**DOI:** 10.1101/2025.08.05.25333011

**Authors:** Asahi Hishida, Yumiko Shirai, Itsuki Kageyama, Sayo Kawai, Tae Sasakabe, Yingsong Lin, Makiko Yamamoto, Michiko Nishio, Hideyuki Okuda, Koji Tanaka, Kazuaki Miyata, Takashi Tamura, Mako Nagayoshi, Takashi Matsunaga, Nobuyuki Hamajima, Kenji Wakai

**Affiliations:** Departments of Public Health, Aichi Medical University, Nagakute, Japan; Departments of Preventive Medicine, Nagoya University Graduate School of Medicine, Nagoya, Japan; Department of Nutrition, Mie University, Tsu, Japan; Department of Clinical Laboratory Tests, Mie University, Tsu, Japan; Department of Surgery, Iga General Hospital, Iga, Mie University, Tsu, Japan; Department of Gastrointestinal and Pediatric Surgery, Mie University, Tsu, Japan; Department of Cardiovascular Medicine, Shiga University of Medical Science, Otsu, Japan

**Keywords:** cohort study, genomic cohort, molecular epidemiology, preventive medicine

## Abstract

We conducted a population-based genome cohort in Iga City, named Iga City Cohort Study, from March 2013 until June 2014, and recruited 1,578 participants in alliance with the J-MICC (Japan Multi-Institutional Collaborative Cohort) Study, under the cooperation of Iga citizens and the support of medical staffs in Iga City General Hospital with the aim of establishing the evidence for personalized disease prevention based on genomic information of each individual. After obtaining informed consent, interviews were conducted using the questionnaire constructed based on those practically used in J-MICC Study, where information on smoking behaviors, drinking habits, physical activities, food intake (food frequency questionnaire, FFQ) and socioeconomic status (SES) including educational backgrounds was asked and collected. Biological samples were collected together with the health check-up examination samples. The samples collected included serum, plasma and buffy coat from which genomic DNA was extracted after centrifuge procedures. In parallel, we performed a public service for *Helicobacter pylori (HP)* eradication to Iga citizens, and research on the minimally invasive diagnostic testing for early detection of cancer is now ongoing. After exclusion of ineligible participants and participants who withdrew, 1,505 subjects remain under follow-up. Effective utilization of the collected data in this cohort study for the health promotion of Iga citizens as well as Japanese citizens is greatly expected.

## Introduction

In Japan, the ageing population is rapidly increasing and chronic diseases such as cardiovascular disease and cancer account for the majority of morbidity and mortality [1]. Therefore, prevention and early detection of these conditions have become an urgent public health priority. Recent progress in the world of medical science, especially the numerous achievements in the field of genetic epidemiology, clarified the possibility of disease prevention tailored to each individual based on genetic information [2].

Iga City, Mie Prefecture, is located in the middle of Japan, between the metropolitan areas of Nagoya and Osaka.

The Iga City Cohort Study is an alliance study of the Japan Multi-Institutional Collaborative Cohort (J-MICC) Study, which is a large-scale nationwide genomic cohort study recruiting approximately 100,000 participants from 14 regions across Japan [3], which is the first large genome cohort study in Japan, with the aim of establishing the evidence for personalized disease prevention based on genomic information of each individual. In this paper, we describe the baseline characteristics of the study participants in the Iga City Cohort Study as the foundation for future follow-up analyses and longitudinal studies on disease incidence and mortality.

## Methods

### Study Participants

We employed a population-based prospective cohort design. Eligible participants were residents of Iga City aged 35–69 years who attended annual health check-ups at the Iga City Medical Health Check-up Center. Individuals unable to provide informed consent or with serious illness were excluded. Among approximately 14,000 residents aged 35–69 years, 1,978 attended the health check-up during the recruitment period, and 1,578 (79.8%) consented to participate in the study. This Iga City Cohort Study is conducted in alliance with the J-MICC Study [3], which recruited about 100,000 participants from the 14 areas of Japan, including the KOPS Study and Kanagawa Cohort Study, which are conducted as alliance studies of the J-MICC Study as well [4]. Written informed consent was obtained from all the participants. The study protocol was approved by the Ethics Committee of Nagoya University Graduate School of Medicine (Approval No. 2012-0138) and the Ethics Committee of Aichi Medical University (Approval No. 2023-167), and written informed consent including permission for follow-up was obtained from all participants.

### Questionnaire

Items included in the questionnaire were constructed based on those practically used in J-MICC Study, the mother cohort study of Iga City Cohort Study. The questionnaire covered smoking status (never, former, current), alcohol consumption (never, occasional, regular), physical activity (leisure-time exercise and daily activity), socioeconomic status (educational attainment, occupation), and dietary habits using a validated food frequency questionnaire (FFQ) consisting of 47 food and beverage items [5,6].

### Biological Samples

Biological samples were collected in parallel with the health check-up examination samples. Practically, the samples collected included serum, plasma and buffy coat. DNA was extracted from buffy coat to investigate genetic susceptibility factors and gene–environment interactions for chronic disease outcomes.

### Statistical Analyses

Differences between the distributions of two continuous variables were examined using Student’s t-test, while differences in categorical variables were assessed using the χ^2^ test. We used Cramér’s V to assess the effect size for associations between categorical variables. Values range from 0 (no association) to 1 (perfect association), with thresholds of 0.10, 0.30, and 0.50 representing small, moderate, and large effects, respectively [7].

## Results

### Study characteristics

The main study characteristics, such as distributions of age or sex, as well as trends in smoking and drinking behaviors of the entire participants of the Iga City Cohort Study are summarized in Table 1. A total of 1,578 participants provided written informed consent, of whom 16 participants were excluded because of lack of eligibility, and 57 subjects withdrew their consent, leaving 1,505 participants eligible for follow-up survey. Elderly participants aged > 60 years old were more frequently observed in males (P = 0.012, χ^2^-test). Both of ever smokers and ever drinkers were more frequently observed in males (P < 0.001 and P < 0.001 by χ^2^-test, respectively). The age distribution of study participants across 5-year age categories showed little difference from that of the overall J-MICC Study cohort (Cramér’s V = 0.057).

**Table 1.** Age distribution, smoking and drinking status of the participants of Iga City Cohort Study.

|  | Male<br>(n = 674) | Female<br>(n = 831) | Total<br>(n = 1,505) |
| --- | --- | --- | --- |
| Age group |  |  |  |
| 35-39 | 109 (16.2%) | 108 (13.0%) | 217 (14.4%) |
| 40-44 | 101 (15.0%) | 132 (15.9%) | 233 (15.5%) |
| 45-49 | 80 (11.9%) | 135 (16.2%) | 215 (14.3%) |
| 50-54 | 117 (17.4%) | 148 (17.8%) | 265 (17.6%) |
| 55-59 | 95 (14.1%) | 141 (17.0%) | 236 (15.7%) |
| 60-64 | 99 (14.7%) | 111 (13.4%) | 210 (14.0%) |
| 65-69 | 73 (10.8%) | 56 ( 6.7%) | 129 ( 8.6%) |
| Smoking |  |  |  |
| Current | 215 (31.9%) | 47 ( 5.7%) | 262 (17.4%) |
| Past | 232 (34.4%) | 54 ( 6.5%) | 286 (19.0%) |
| Never | 180 (26.7%) | 632 (76.1%) | 812 (54.0%) |
| (unknown) | 47 ( 7.0%) | 98 (11.8%) | 145 ( 9.6%) |
| Drinking |  |  |  |
| Current | 498 (73.9%) | 286 (34.4%) | 784 (52.1%) |
| Past | 19 ( 2.8%) | 13 ( 1.6%) | 32 ( 2.1%) |
| Never | 156 (23.1%) | 524 (63.1%) | 680 (45.2%) |
| (refused) | 1 ( 0.1%) | 6 ( 0.7%) | 7 ( 0.5%) |
| (unknown) | 0 ( 0.0%) | 2 ( 0.2%) | 2 ( 0.1%) |

Smoking and alcohol consumption data were unavailable for 7 participants (0.5%) because of an irretrievable data entry error at baseline.

### Number of participants and participation rates by time

Number of participants and participation rates by chronological time (month) are as shown in Figure 1. Notably, the average participation rate exceeded 80%.

**Fig. 1.**
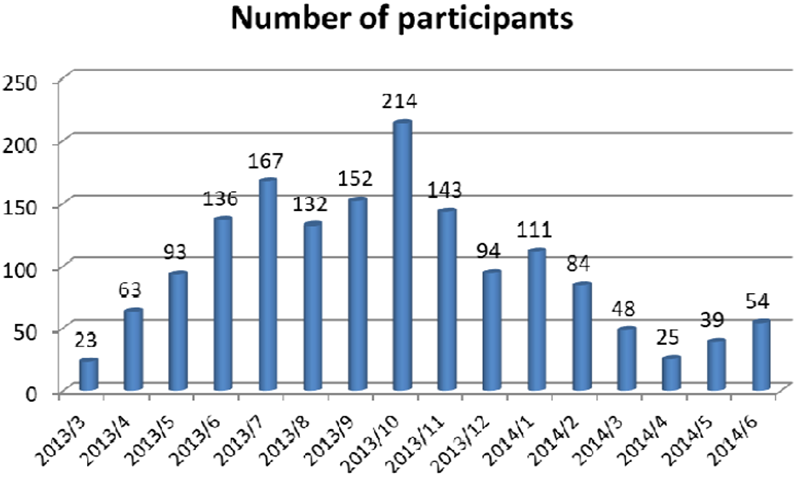
Number of participants by month. Values shown in this figure are aggregate data based on the baseline survey at the time of enrollment.

**Fig. 2.**
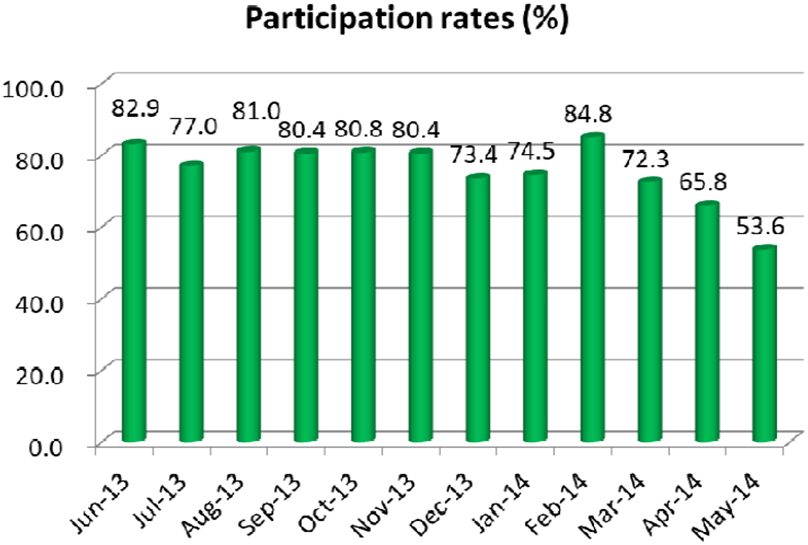
Participation rate by month. Data were not available for months other than the months presented. Values shown in this figure are aggregate data based on the baseline survey at the time of enrollment.

### Anthropometric and laboratory data

Representative anthropometric measures and laboratory data of the participants at baseline are described in Table 2. Blood pressures (BPs) (both systolic and diastolic) were higher in males, high-density lipoprotein cholesterols (HDL-Cs) were higher in females, and liver enzymes (AST: aspartate aminotransferase; ALT: alanine aminotransferase; GGT: γ-glutamyl transferase) were higher in males (P < 0.001, P < 0.001, and P < 0.001, by Student’s t-test).

**Table 2.**
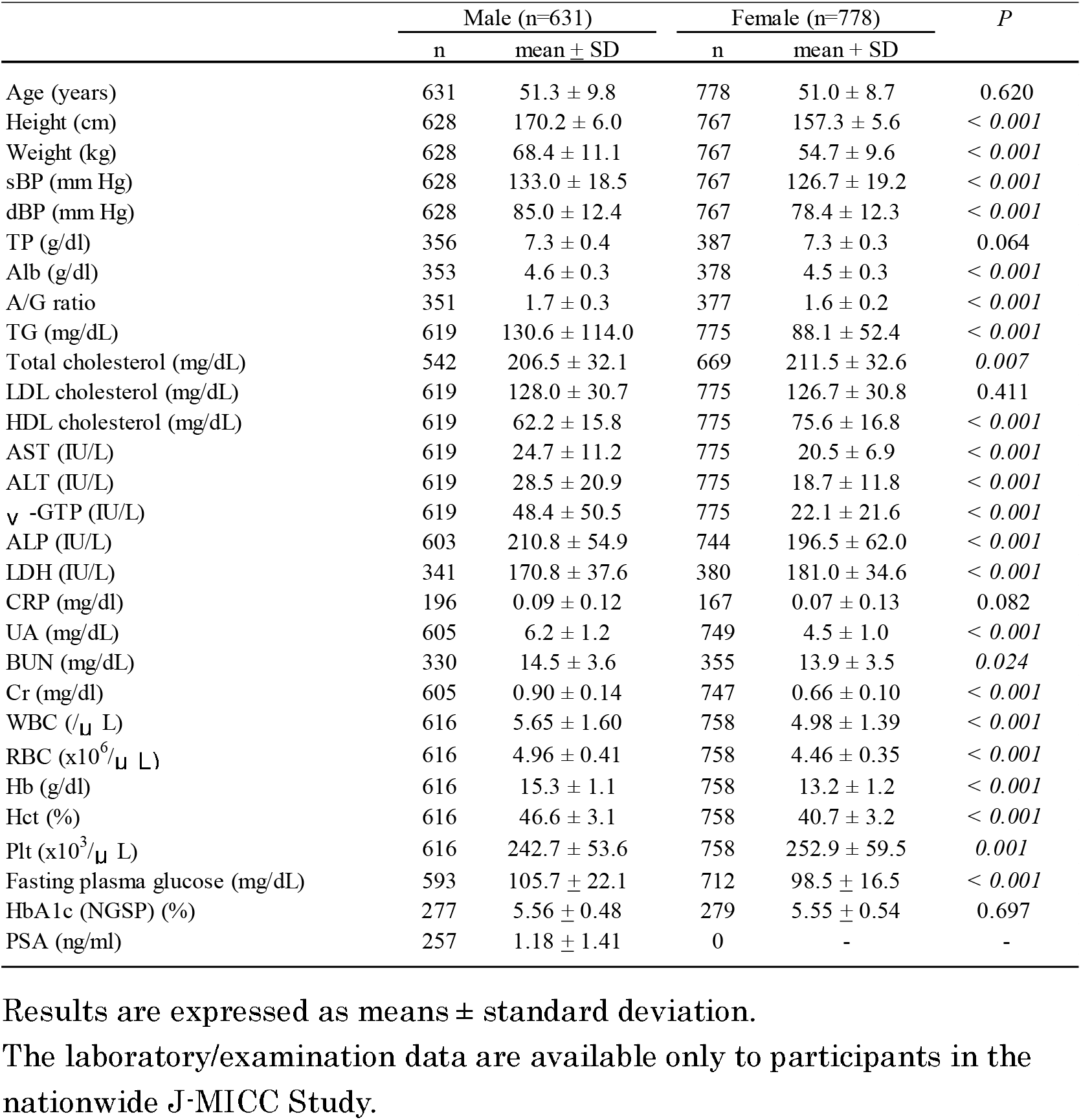
Laboratory data of the study participants.

|  | Male (n=631) |  | Female (n=778) |  | P |
| --- | --- | --- | --- | --- | --- |
| | n | mean ± SD | n | mean ± SD | |
| Age (years) | 631 | 51.3 ± 9.8 | 778 | 51.0 ± 8.7 | 0.620 |
| Height (cm) | 628 | 170.2 ± 6.0 | 767 | 157.3 ± 5.6 | < 0.001 |
| Weight (kg) | 628 | 68.4 ± 11.1 | 767 | 54.7 ± 9.6 | < 0.001 |
| sBP (mm Hg) | 628 | 133.0 ± 18.5 | 767 | 126.7 ± 19.2 | < 0.001 |
| dBp (mm Hg) | 628 | 85.0 ± 12.4 | 767 | 78.4 ± 12.3 | < 0.001 |
| TP (g/dl) | 356 | 7.3 ± 0.4 | 387 | 7.3 ± 0.3 | 0.064 |
| Alb (g/dl) | 353 | 4.6 ± 0.3 | 378 | 4.5 ± 0.3 | < 0.001 |
| A/G ratio | 351 | 1.7 ± 0.3 | 377 | 1.6 ± 0.2 | < 0.001 |
| TG (mg/dL) | 619 | 130.6 ± 114.0 | 775 | 88.1 ± 52.4 | < 0.001 |
| Total cholesterol (mg/dL) | 542 | 206.5 ± 32.1 | 669 | 211.5 ± 32.6 | 0.007 |
| LDL cholesterol (mg/dL) | 619 | 128.0 ± 30.7 | 775 | 126.7 ± 30.8 | 0.411 |
| HDL cholesterol (mg/dL) | 619 | 62.2 ± 15.8 | 775 | 75.6 ± 16.8 | < 0.001 |
| AST (IU/L) | 619 | 24.7 ± 11.2 | 775 | 20.5 ± 6.9 | < 0.001 |
| ALT (IU/L) | 619 | 28.5 ± 20.9 | 775 | 18.7 ± 11.8 | < 0.001 |
| γ-GTP (IU/L) | 619 | 48.4 ± 50.5 | 775 | 22.1 ± 21.6 | < 0.001 |
| ALP (IU/L) | 603 | 210.8 ± 54.9 | 744 | 196.5 ± 62.0 | < 0.001 |
| LDH (IU/L) | 341 | 170.8 ± 37.6 | 380 | 181.0 ± 34.6 | < 0.001 |
| CRP (mg/dl) | 196 | 0.09 ± 0.12 | 167 | 0.07 ± 0.13 | 0.082 |
| UA (mg/dL) | 605 | 6.2 ± 1.2 | 749 | 4.5 ± 1.0 | < 0.001 |
| BUN (mg/dL) | 330 | 14.5 ± 3.6 | 355 | 13.9 ± 3.5 | 0.024 |
| Cr (mg/dl) | 605 | 0.90 ± 0.14 | 747 | 0.66 ± 0.10 | < 0.001 |
| WBC (/$\mu$ L) | 616 | 5.65 ± 1.60 | 758 | 4.98 ± 1.39 | < 0.001 |
| RBC ($\times 10^6$/$\mu$ L) | 616 | 4.96 ± 0.41 | 758 | 4.46 ± 0.35 | < 0.001 |
| Hb (g/dl) | 616 | 15.3 ± 1.1 | 758 | 13.2 ± 1.2 | < 0.001 |
| Hct (%) | 616 | 46.6 ± 3.1 | 758 | 40.7 ± 3.2 | < 0.001 |
| Plt ($\times 10^3$/$\mu$ L) | 616 | 242.7 ± 53.6 | 758 | 252.9 ± 59.5 | 0.001 |
| Fasting plasma glucose (mg/dL) | 593 | 105.7 ± 22.1 | 712 | 98.5 ± 16.5 | < 0.001 |
| HbA1c (NGSP) (%) | 277 | 5.56 ± 0.48 | 279 | 5.55 ± 0.54 | 0.697 |
| PSA (ng/ml) | 257 | 1.18 ± 1.41 | 0 | - | - |

Results are expressed as means ± standard deviation.
The laboratory/examination data are available only to participants in the nationwide J-MICC Study.

### Other representative data obtained by questionnaire

Frequencies of staple food intake as the representative data for the food frequency questionnaire, and the results of the questionnaire on *Helicobacter pylori (HP)* infection of the participants, which is a unique question for Iga City Cohort Study, are described in Table 3 and Table 4. The proportion of participants who had not undergone testing for *HP* infections was not significantly different between males and females (71.4% vs. 68.5%, P = 0.224 by by χ^2^-test), whereas the proportion of participants who tested positive for *HP* infections was slightly higher in males than in females, although the difference did not reach statistical significance (P = 0.057 by χ^2^-test). For the staple food intake, participants who take white rice for breakfast were more frequent in males than in females (P < 0.001 by χ^2^-test), whereas those who have breads for breakfast more than one day/week were more frequent in females (P < 0.001 by χ^2^-test).

**Table 3.** Summary data for the questionnaire on macronutrient intake of the study participants.

|  | rarely | 1-3 a month | 1-2 a week | 3-4 a week | 5-6 a week | everyday | others | Total | P |
| --- | --- | --- | --- | --- | --- | --- | --- | --- | --- |
| Breakfast |  |  |  |  |  |  |  |  |  |
| White rice |  |  |  |  |  |  |  |  |  |
| Males | 252 (37.4%) | 24 ( 3.6%) | 40 ( 5.9%) | 36 ( 5.3%) | 80 (11.9%) | 148 (22.0%) | 94 (13.9%) | 674 | < 0.001 |
| Females | 379 (45.6%) | 50 ( 6.0%) | 77 ( 9.3%) | 70 ( 8.4%) | 90 (10.8%) | 119 (14.3%) | 46 (5.5%) | 831 |  |
| Bread |  |  |  |  |  |  |  |  |  |
| Males | 191 (28.3%) | 35 ( 5.2%) | 107 (15.9%) | 39 ( 5.8%) | 51 ( 7.6%) | 164 (24.3%) | 87 (12.9%) | 674 | < 0.001 |
| Females | 138 (16.6%) | 57 ( 6.9%) | 119 (14.3%) | 87 (10.5%) | 87 (10.5%) | 302 (36.3%) | 41 (4.9%) | 831 |  |
| Noodle |  |  |  |  |  |  |  |  |  |
| Males | 504 (74.8%) | 20 ( 3.0%) | 23 ( 3.4%) | 0 (0.0%) | 0 (0.0%) | 1 ( 0.1%) | 126 (18.7%) | 674 | < 0.001 |
| Females | 725 (87.2%) | 23 ( 2.8%) | 11 ( 1.3%) | 0 (0.0%) | 0 (0.0%) | 1 ( 0.1%) | 71 (8.5%) | 831 |  |
| Lunch |  |  |  |  |  |  |  |  |  |
| White rice |  |  |  |  |  |  |  |  |  |
| Males | 22 ( 3.3%) | 8 ( 1.2%) | 44 ( 6.5%) | 72 (10.7%) | 161 (23.9%) | 296 (43.9%) | 71 (10.5%) | 674 | < 0.001 |
| Females | 53 ( 6.4%) | 15 ( 1.8%) | 68 ( 8.2%) | 153 (18.4%) | 267 (32.1%) | 253 (30.4%) | 22 (2.6%) | 831 |  |
| Bread |  |  |  |  |  |  |  |  |  |
| Males | 420 (62.3%) | 53 ( 7.9%) | 60 ( 8.9%) | 17 ( 2.5%) | 10 ( 1.5%) | 5 ( 0.7%) | 109 (16.2%) | 674 | < 0.001 |
| Females | 445 (53.5%) | 113 (13.6%) | 165 (19.9%) | 40 ( 4.8%) | 3 ( 0.4%) | 11 ( 1.3%) | 54 (6.5%) | 831 |  |
| Noodle |  |  |  |  |  |  |  |  |  |
| Males | 272 (40.4%) | 109 (16.2%) | 138 (20.5%) | 51 ( 7.6%) | 11 ( 1.6%) | 5 ( 0.7%) | 87 (12.9%) | 674 | < 0.001 |
| Females | 319 (38.4%) | 161 (19.4%) | 262 (31.5%) | 39 ( 4.7%) | 14 ( 1.7%) | 1 ( 0.1%) | 35 (4.2%) | 831 |  |
| Dinner |  |  |  |  |  |  |  |  |  |
| White rice |  |  |  |  |  |  |  |  |  |
| Males | 40 ( 5.9%) | 16 ( 2.4%) | 26 ( 3.9%) | 41 ( 6.1%) | 117 (17.4%) | 343 (50.9%) | 91 (13.5%) | 674 | < 0.001 |
| Females | 40 ( 4.8%) | 14 ( 1.7%) | 41 ( 4.9%) | 70 ( 8.4%) | 161 (19.4%) | 456 (54.9%) | 49 (5.9%) | 831 |  |
| Bread |  |  |  |  |  |  |  |  |  |
| Males | 523 (77.6%) | 10 ( 1.5%) | 11 ( 1.6%) | 1 ( 0.1%) | 0 (0.0%) | 0 (0.0%) | 129 (19.1%) | 674 | < 0.001 |
| Females | 718 (86.4%) | 23 ( 2.8%) | 13 ( 1.6%) | 7 ( 0.8%) | 0 (0.0%) | 0 (0.0%) | 70 (8.4%) | 831 |  |
| Noodle |  |  |  |  |  |  |  |  |  |
| Males | 312 (46.3%) | 120 (17.8%) | 111 (16.5%) | 19 ( 2.8%) | 2 ( 0.3%) | 3 ( 0.4%) | 107 (15.9%) | 674 | < 0.001 |
| Females | 418 (50.3%) | 193 (23.2%) | 144 (17.3%) | 21 ( 2.5%) | 2 ( 0.2%) | 1 ( 0.1%) | 52 (6.3%) | 831 |  |

\*Others: no answer, multiple answers, unknown.

**Table 4.** Summary data for the questionnaire on Helicobacter pylori infection of the study participants.

| *HP* infection | Male<br>(n = 674) | Female<br>(n = 831) | Total<br>(n = 1,505) |
| --- | --- | --- | --- |
| No test | 481 (71.4%) | 569 (68.5%) | 1,050 (69.8%) |
| Test negative | 58 ( 8.6%) | 131 (15.8%) | 189 (12.6%) |
| Positive, no eradication | 11 ( 1.6%) | 21 ( 2.5%) | 32 ( 2.1%) |
| Positive, eradicated | 58 ( 8.6%) | 41 ( 4.9%) | 99 ( 6.6%) |
| Eradication failure | 4 ( 0.6%) | 3 ( 0.4%) | 7 ( 0.5%) |
| No test after eradication | 26 ( 3.9%) | 49 ( 5.9%) | 75 ( 5.0%) |
| No recollection | 35 ( 5.2%) | 14 ( 1.7%) | 49 ( 3.3%) |
| Refused | 1 ( 0.1%) | 1 ( 0.1%) | 2 ( 0.1%) |
| Unknown | 0 ( 0.0%) | 2 ( 0.2%) | 2 ( 0.1%) |

*HP: Helicobacter pylori.*

## Discussion

The Iga City Cohort Study achieved a high participation rate (approximately 80%), which is higher than that of many other Japanese population-based cohorts. This success likely reflects strong collaboration with the municipal health center and contributed substantially to the overall goal of recruiting 100,000 participants in the parent study, the J-MICC Study [3]. We also observed clear sex differences in blood pressure, liver enzyme levels, and *Helicobacter pylori (HP)* infection rates, which will be important for future analyses linking baseline characteristics with disease incidence. The proportion of female participants was slightly higher than that of males, reflecting the usual trend among examinees at the Iga City Health Check-up Center, where breast and gynecological examinations for women are emphasized.

Iga City is located in the center of the Iga Basin, within the mountainous Kii Peninsula of Japan. Due to its distance from major metropolitan areas such as Osaka, Nagoya, and even the prefectural capital of Tsu (more than an hour by car), the city has long experienced a shortage of full-time medical doctors. This situation, combined with lower motivation among citizens to undergo regular health check-ups, may contribute to delayed diagnosis of life-threatening diseases, including cancers and cardiovascular diseases. Consequently, emergency rooms in Iga hospitals are frequently busy, and outpatient clinics for cancer palliative care play a crucial role in local medical services.

Despite these challenges, the medical staff of Iga City General Hospital, including physicians, nurses, nutritionists, and allied health professionals, have been working actively to improve the situation. With the support of the hospital director, who has a strong academic interest in medical research, we were able to complete the baseline survey successfully. The hospital continues to provide palliative care, including nutritional interventions, for patients with gastrointestinal cancers through its “Cancer Immuno-nutrition Support Center,” established in 2011. This effort has led to the accumulation of substantial clinical data, contributing to evidence-based palliative care. Examples include the demonstrated benefits of fish oil administration for advanced gastrointestinal cancer patients with inflammation [8], the influence of host genetic factors on cancer survival [9,10], the utility of the modified Glasgow Prognostic Scale (mGPS) in colorectal cancer prognosis [11], and the potential clinical relevance of elevated alanine aminotransferase (ALT), total bilirubin (T-bil), blood urea nitrogen (BUN), creatinine (Cr), and decreased platelet counts as markers of mortality in terminal cancer patients [12].

A major challenge that remains is the low health check-up rate among Iga citizens, which may reduce survival rates by delaying diagnoses. Effective countermeasures could include community education on the importance of early and regular check-ups. We also implemented a public service program for *HP* eradication among citizens aged 20–69 years to reduce HP infection rates and related healthcare costs. A related analysis of post-eradication cost reduction was previously presented at a scientific meeting [13]. Another future priority may be the wider adoption of minimally invasive diagnostic testing for early cancer detection [14,15]. In parallel, the Iga City Cohort Study is expected to contribute to the identification of gene–environment interactions for cancer prevention, complementing the ongoing activities of its parent cohort, the J-MICC Study [16–19].

Participant communication has often been underemphasized in traditional cohort research, where primary attention has generally been directed toward study design, recruitment, and data collection. Nevertheless, for future community-based biomarker cohorts, more structured and participant-centered communication processes may be valuable for sustaining trust and supporting long-term research continuity. Given that the age distribution of the Iga cohort closely mirrors that of the overall J-MICC cohort, suggesting its value as a representative regional population in Japan, continued sincere efforts are needed to honor the goodwill of community participants. Sustaining public trust will require thoughtful engagement and clear communication throughout future research initiatives.

In summary, we conducted the baseline survey of the Iga City Cohort Study from March 2013 until June 2014, and recruited 1,578 participants with lifestyle data and biospecimens including serum, plasma and genomic samples, with the cooperation of Iga citizens and the support of medical staff in Iga City General Hospital, of whom 1,505 participants remain under follow-up. Effective utilization of the collected data in this cohort study for the health promotion of Iga citizens as well as Japanese citizens is greatly expected.

## Data Availability

All data produced in the present study are available upon reasonable request to the authors

## ACKNOWLEDGEMENTS

The authors are grateful to Ms. Yoko Mitsuda, Ms. Rie Terasawa and Ms. Keiko Shibata for their technical assistance.

## FUNDING

This study was supported by Grants-in-Aid for Scientific Research for Priority Areas of Cancer (No. 17015018) and Innovative Areas (No. 221S0001) and by the Japan Society for the Promotion of Science (JSPS) KAKENHI Grant (No. 16H06277 [CoBiA]) from the Japanese Ministry of Education, Culture, Sports, Science and Technology.

We declare that all the authors approved the publication of this study paper.

